# Beyond the freedom to refuse patient: a retrospective comparative study of emergency transportation during the COVID-19 pandemic in Japan

**DOI:** 10.1101/2025.08.19.25334036

**Authors:** Kaiho Hirata, Takuyo Chiba, Reo Takaku, Chen Meilai, Shunya Ikeda, Takashi Shiga

## Abstract

Emergency room overcrowding and ambulance diversion have been a significant problem worldwide and it became more apparent during the COVID-19 pandemic. In Japan, the Dedicated Emergency Physician (DEP) model has been associated with reduced transport time, but it is unclear whether this benefit persisted during the pandemic. We analyzed changes transport outcomes during the COVID-19 pandemic in Japanese regions with and without DEP hospitals in order to evaluate the effectiveness of the DEP model and to identify factors which could improve transport outcomes. Using nationwide data from January 2015 to December 2021, we analyzed three target areas: Urayasu-Ichikawa (DEP Group 1), Shonan-Fujisawa (DEP Group 2), and Ichinomiya-Toyota (Non-DEP Group 3). DEP Groups 1 and 2 contained DEP hospitals, while Non-DEP Group 3 was selected for comparable population size and strong pre-pandemic transport performance. To minimize the impact of regional variations in COVID-19 prevalence, we compared the changes in transport outcomes before and after the pandemic between the target areas and the nearby comparison areas. In total, there were 150,856 transports in Group 1, 186,965 in Group 2, and 516,655 in Group 3. In the target areas of Groups 2 and 3, transport time changes were significantly shorter by 2.016 and 0.606 minutes, respectively, compared with comparison areas. Moreover, these areas had significantly lower odds of transportation difficulty (Group 2: OR 0.131, 95% CI 0.110–0.157; Group 3: OR 0.086, 95% CI 0.066–0.112). We found that common characteristics of these areas were densely located large-scale hospitals and makeshift buildings for COVID-19 patients just next to large-scale hospitals. These findings suggest that DEP hospitals alone did not guarantee favorable transport outcomes during the pandemic. A sufficient number of large-scale hospitals and nearby temporary facilities may be crucial to maintaining effective emergency transport under pandemic conditions.

## Introduction

Emergency department (ED) overcrowding and ambulance diversion have long been recognized as important problems that lead to increased morbidity and mortality^1–3^, and which will potentially worsen for decades owing to rapid aging in developed countries. Although several studies have proposed solutions, these problems have never been resolved now^4–6^. Furthermore, during the COVID-19 pandemic, ED overcrowding and ambulance diversion became more apparent, mainly because of the increased rate of patients with high-acuity conditions and patient management changes, leading to prolonged lengths of stay in the ED^7–9^.

This problem has been an important issue in Japan for several decades. Transportation time in Japan has become longer, and one of the reasons is the unique emergency medical service (EMS) system, where each hospital has the discretion to decide whether to accept or reject an EMS request for patient transportation^10^. Based on this “freedom to accept/reject” system, the EMS rejection by hospital is a widespread phenomenon in Japan. Furthermore, the number of difficult hospital acceptance cases, defined as EMS rejections of more than three times, has increased dramatically since the onset of the COVID-19 outbreak in 2020, according to the Ministry of Health, Labour and Welfare^11^. In 2022, a quarter of severely ill patients were rejected more than once when they called an ambulance, with the maximum number of rejections being 272^12^.

The number of EMS rejections was low in areas with a Dedicated Emergency Physician (DEP) Model and was reduced when the government increased reimbursement for emergency care^13, 14^. In particular, DEP Model, which is similar to the emergency room (ER) system in the U.S., was shown to have a great impact on pre-hospital transportation time, reducing it by approximately 7 minutes^13^. However, little is known about whether this model contributed to positive effects during the COVID-19 pandemic and what factors could improve transport outcomes.

To address this knowledge gap, we present a retrospective study examining changes in transportation outcomes before and after COVID-19 pandemic in areas with and without DEP model. We used comprehensive emergency transport data from January 2015 to December 2021. We aimed to examine whether the DEP system remains associated with favorable outcomes and whether other factors contributed to favorable outcomes.

## Materials and methods

### Study Design

This was a retrospective observational study based on a Japanese national database from January 1, 2015, to December 31, 2021. All patients who were transported to hospitals by ambulance in Japan, except for Tokyo, were registered. The database includes time course of transportation, age, gender, location of call, transport reason, and severity (death, severe, moderate and mild). The severity was determined by the physician in charge at the time of hospital arrival. Severe and moderate patients are those who expected to be hospitalized for three weeks or more and for less than three weeks, respectively. Mild patients are those who expected not to require hospitalization.

We selected two regions as DEP model target areas based on the previous research^13^ (DEP Group 1: Urayasu city and Ichikawa city in Chiba prefecture; and DEP Group 2: Kamakura city, Chigasaki city, Fujisawa city and Zushi city in Kanagawa prefecture). These areas are known for the presence of DEP hospitals (DEP Group 1: Tokyo Bay Urayasu Ichikawa Medical Center; and DEP Group 2: Shonan Kamakura General Hospital), which is defined as hospitals which have more than 15 DEP physicians including senior staff and senior residents. Among the five major metropolitan areas in Japan, Aichi Prefecture (population of 7.4 million) was selected for its comparable scale— similar to Chiba Prefecture (population of 6.2 million) and Kanagawa Prefecture (population of 9.2 million)—and because it also faces the sea. Within Aichi Prefecture, we selected one region as a Non-DEP model target area based on its favorable emergency transport performance before the COVID-19 pandemic (Non-DEP Group 3: Ichinomiya city, Seto city, Kasugai city, Tsushima city, Toyota city and Inuyama city in Aichi prefecture).

For each area, we selected a nearby area for comparison that was geographically adjacent to the selected areas and comparable in terms of population, size, and location. The comparison areas were Funabashi City for DEP Group 1; Odawara City, Isehara City, Hatano City, and Sagamihara City for DEP Group 2; and Nagoya City for Non-DEP Group 3. The geographical relationship between the target and comparison areas is shown in S1 Figure. Patients with missing data and patients with transportation reasons other than medical illnesses were excluded from analysis.

In Chiba Prefecture, which has a population of 6.2 million, one of the top five hospitals accepting emergency transport received about 7,000 patients. Therefore, we defined a large-scale hospital as one that accepts over 7,000 ambulances per year and we calculated Herfindahl-Hirschman Index of transportations to large-scale hospitals in each area. This was calculated based on the Hospital Function Report in 2022^15^.

This study was approved by the Ethics Committee of the International University of Health and Welfare, Narita Hospital (24-Im-014 June 25, 2024). We accessed the data for research purposes on July 1 2024. Informed consent was not required to conduct this study because it was an observational study without intervention, and the data did not include identifiable personal information.

### Study Outcomes

The primary outcomes were transportation time, defined as time from arrival at the scene to arrival at the hospital and the number of the transportation difficulty cases, defined as a case with ≥4 phone calls by EMS to hospitals until the patient is accepted^16^.

### Statistical Analysis

To minimize regional disparities in COVID-19 spread, we compared changes in transportation performance between target and comparison areas in each group based on the assumption that geographically adjacent regions with similar population sizes and characteristics would be affected similarly by COVID-19 pandemic. This approach likely reduced, though may not have entirely eliminated, bias due to regional variation in pandemic impact.

We conducted a multivariate analysis to compare the changes in transportation time and the number of transportation difficulty cases before and after the COVID-19 pandemic in the target and comparison areas. These changes were evaluated using an interaction term of a binary variable for the target areas and a binary variable for the COVID-19 period, similar to a difference-in-difference analysis. The COVID-19 period was defined as the period from January 2020, when the first COVID-19 outbreak occurred in Japan, to December 2021.

In the multivariate analysis, we controlled for the time from emergency call to arrival at the scene, number of emergency calls within the same hour at the same EMS department, location of call, severity (death, severe, and moderate), and basic patient characteristics such as age and sex. The fixed effects for the EMS department and years/month were also controlled. To address the serial correlation within each EMS department, standard errors were clustered at EMS department level^17^. We also conducted a stratified analysis of transportation time and difficulty. Stratification factors included respiratory disease, oxygenation, and severity (severe, moderate, and mild).

Since prehospital transportation time was a continuous variable, we used ordinary least squares to implement a multivariate analysis. Similarly, we used logistic regression analysis for transportation difficulty because it is a binary variable. For each outcome, we also performed stratified analyses based on following categories: whether the primary reason for transport was respiratory diseases, oxygen supply during transport, and the severity (severe, moderate, and mild). Stata (version 18.0; Stat Corp, College Station, TX, USA) was used for statistical analysis, and a two-tailed P value of < 0.05 was considered statistically significant.

## Results

A total of 19,953,584 patients were included in the study. After patients outside the target and comparison areas, patients with incomplete data or transportation reasons other than medical illnesses were excluded, 150,856 patients in Group 1; 186,965 in Group 2; and 516,656 in Group 3 were included in the analysis (Fig 1).

**Fig 1:**
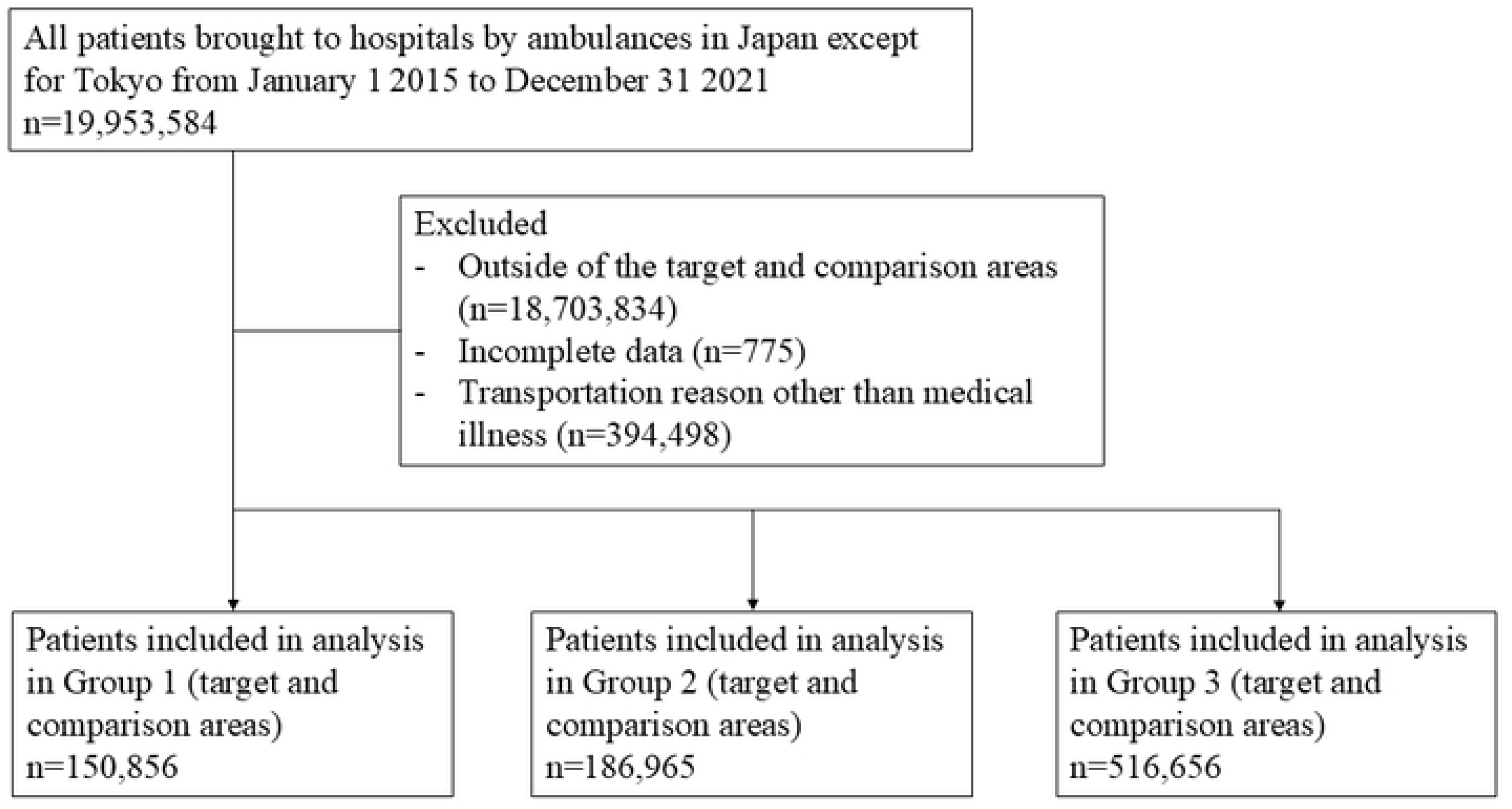
Patient enrolment.

The background information of each group is summarized in Table 1. The percentage of transportation to large-scale hospitals (hospitals accepting over 7,000 ambulances per year) was much higher in the target areas of Groups 2 and 3 than in Group 1. The baseline characteristics of patients in each group are shown in Table 2. The mean time from call to hospital was shortest in Group 3 target area, and the percentage of transportation difficulty cases was lowest in Group 3 target area.

**Table 1.** Background characteristics of each group.

|  | Group 1 - Chiba |  | Group 2 - Kanagawa |  | Group 3 - Aichi |  |
| --- | --- | --- | --- | --- | --- | --- |
|  | Target area | Comparison area | Target area | Comparison area | Target area | Comparison area |
| Hospitals accepting ambulances | 11 | 9 | 17 | 14 | 21 | 52 |
| Large-scale hospitals | 1 | 0 | 3 | 0 | 8 | 6 |
| Annual transportations | 32913 | 23146 | 48127 | 34777 | 78730 | 120346 |
| Annual transportations per hospitals accepting ambulances | 2992 | 2572 | 2831 | 2484 | 3749 | 2314 |
| The percentage of transportations to large-scale hospitals in whole transportations (%) | 31 | 0 | 68 | 0 | 80 | 46 |
| Herfindahl-Hirschman Index of transportations to large-scale hospitals | 1956 | 1467 | 1782 | 1213 | 904 | 537 |
| Population | 668038 | 642907 | 909064 | 1178568 | 1642196 | 2332176 |
| Percentage of people older than 65 (%) | 21 | 24 | 27 | 28 | 26 | 25 |

Note: The difinition of large-scale hospital is a hospital which accepts more than 7,000 ambulances
per year.

**Table 2.** Patient characteristics.

|  | Group 1 - Chiba |  | Group 2 - Kanagawa |  | Group 3 - Aichi |  |
| --- | --- | --- | --- | --- | --- | --- |
|  | Target area | Comparison area | Target area | Comparison area | Target area | Comparison area |
| **Observations** | 73,481 | 77,375 | 118,274 | 68,691 | 197,920 | 318,736 |
| **Time from emergency call to the hospital, mean (min)** | 41 | 46 | 33 | 40 | 29 | 31 |
| **Time from emergency call to the scene, mean (min)** | 8 | 9 | 7 | 8 | 7 | 6 |
| **Number of calls within the same hour** | 2 | 3 | 2 | 2 | 2 | 11 |
| **Age, median (IQR)** | 67(40-81) | 72(47-83) | 75(52-84) | 74(53-83) | 74(52-83) | 71(47-83) |
| **Age group, n (%)** |  |  |  |  |  |  |
| 0s | 4,454(6.06) | 3,832(4.95) | 5,624(4.76) | 2,695(3.92) | 8,918(4.51) | 13,525(4.24) |
| 10s | 2,366(3.22) | 2,080(2.69) | 3,115(2.63) | 1,937(2.82) | 5,609(2.83) | 8,421(2.64) |
| 20s | 6,148(8.37) | 5,068(6.55) | 5,554(4.70) | 3,363(4.90) | 8,610(4.35) | 23,489(7.37) |
| 30s | 5,256(7.15) | 4,251(5.49) | 5,238(4.43) | 2,957(4.30) | 8,848(4.47) | 17,706(5.56) |
| 40s | 6,115(8.32) | 5,665(7.32) | 7,735(6.54) | 4,439(6.46) | 13,156(6.65) | 23,374(7.33) |
| 50s | 6,980(9.50) | 6,329(8.18) | 9,567(8.09) | 5,594(8.14) | 15,517(7.84) | 27,735(8.70) |
| 60s | 7,815(10.64) | 7,439(9.61) | 11,281(9.54) | 7,431(10.82) | 20,814(10.52) | 34,337(10.77) |
| 70s | 14,104(19.19) | 17,143(22.16) | 23,925(20.23) | 15,654(22.79) | 46,957(23.73) | 65,176(20.45) |
| 80s | 15,148(20.61) | 19,103(24.60) | 33,246(28.11) | 17,842(25.97) | 52,434(26.49) | 78,298(24.57) |
| 90+ | 5,095(6.93) | 6,465(8.36) | 12,989(10.98) | 6,779(9.87) | 17,057(8.62) | 26,675(8.37) |
| **Female sex, n (%)** | 36,251(49.33) | 38,399(49.63) | 59,967(50.70) | 33,149(48.26) | 93,602(47.29) | 152,823(47.95) |
| **Location, n (%)** |  |  |  |  |  |  |
| Home | 54,604(74.31) | 57,033(73.71) | 88,641(74.95) | 51,060(74.33) | 149,540(75.56) | 224,602(70.47) |
| **Transport reason, n (%)** |  |  |  |  |  |  |
| Brain disease | 3,625(4.93) | 3,978(5.14) | 4,679(3.96) | 4,405(6.41) | 11,928(6.03) | 17,377(5.45) |
| Cardiovascular disease | 3,540(4.82) | 4,472(5.78) | 9,896(8.37) | 4,686(6.82) | 16,054(8.11) | 24,697(7.75) |
| Gastrointestinal disease | 6,015(8.19) | 5,547(7.17) | 7,628(6.45) | 4,427(6.44) | 14,541(7.35) | 32,291(10.13) |
| Respiratory disease | 6,029(8.20) | 7,425(9.60) | 8,331(7.04) | 5,309(7.73) | 17,326(8.75) | 33,310(10.45) |
| Psychiatric disease | 1,528(2.08) | 2,168(2.80) | 2,688(2.27) | 1,805(2.63) | 5,024(2.54) | 8,852(2.78) |
| Neurological disease | 1,329(1.81) | 1,009(1.30) | 3,018(2.55) | 1,934(2.82) | 11,057(5.59) | 12,311(3.86) |
| Urologic disease | 1,991(2.71) | 2,031(2.62) | 2,977(2.52) | 1,918(2.79) | 7,170(3.62) | 10,877(3.41) |
| Neoplastic disease | 591(0.80) | 1,391(1.80) | 952(0.80) | 992(1.44) | 3,434(1.74) | 4,406(1.38) |
| Others | 29,504(40.15) | 8,263(10.68) | 47,504(40.16) | 9,265(13.49) | 27,719(14.01) | 35,689(11.20) |
| Unclear | 19,329(26.30) | 41,091(53.11) | 30,601(25.87) | 33,950(49.42) | 83,667(42.27) | 138,926(43.59) |
| **Oxygen supply during transportation, n(%)** | 11,024(15.00) | 12,673(16.38) | 23,774(20.10) | 15,266(22.22) | 43,966(22.21) | 64,062(20.10) |
| **Severity, n (%)** |  |  |  |  |  |  |
| Death | 419(0.57) | 398(0.51) | 2,060(1.74) | 1,543(2.25) | 3,858(1.95) | 1,470(0.46) |
| Severe | 4,460(6.07) | 4,810(6.22) | 8,946(7.56) | 5,975(8.55) | 14,728(7.44) | 10,963(3.44) |
| Moderate | 32,842(44.69) | 36,436(47.09) | 66,246(56.01) | 34,801(50.66) | 84,586(42.74) | 124,704(39.12) |
| Mild | 35,760(48.67) | 35,731(46.18) | 41,022(34.68) | 26,472(38.54) | 94,748(47.87) | 181,599(56.97) |
| **Transportation difficulty cases, n (%)** | 2,753(3.75) | 3,563(4.60) | 181(0.15) | 2,206(3.21) | 121(0.06) | 4,842(1.52) |

The raw data trends for the transportation time are shown in Fig 2. Graphically, the transportation time increased in the target area in Group 1 in a manner similar to that in the comparison area. However, although the comparison area showed an increasing trend, the target area showed an unchanged trend during the pandemic in Group 2. In Group 3, there was an increasing trend in the target area; however, it was less prominent than that in the comparison area. Fig 3 shows the trends in the number of transportation-difficulty case. Although the target area in Group 1 showed an increasing trend similar to that of the comparison area, the target areas in Groups 2 and 3 showed unchanged trends during the pandemic. The results of the multivariate regression and stratified analyses of transportation time and transportation difficulty changes are summarized in Table 3. In Groups 2 and 3 target areas, the relative changes in transportation time compared with the comparison areas were −2.016 min (95% CI −3.631 to −0.400) and −0.606 min (95% CI −1.114 to - 0.098), respectively. The odds ratio of transportation difficulty in the target areas compared to the comparison areas in Groups 2 and 3 were 0.131 (95% CI 0.010–0.157) and 0.086 (95% CI 0.066– 0.112), respectively. This result was observed even after stratification by patients with respiratory diseases and patients with a demand for oxygen. In contrast, there was a significantly higher increase in the odds ratio of transportation difficulty in the target area than in the comparison area in Group 1 (odds ratio 1.393, 95% CI 1.314–1.477). This unfavorable change was more prominent in patients with respiratory diseases (odds ratio 2.767, 95% CI 2.349–3.260). The full estimation results with coefficients (odds ratios) and confidence intervals of all covariates are reported in S2 Table.

**Fig 2:**
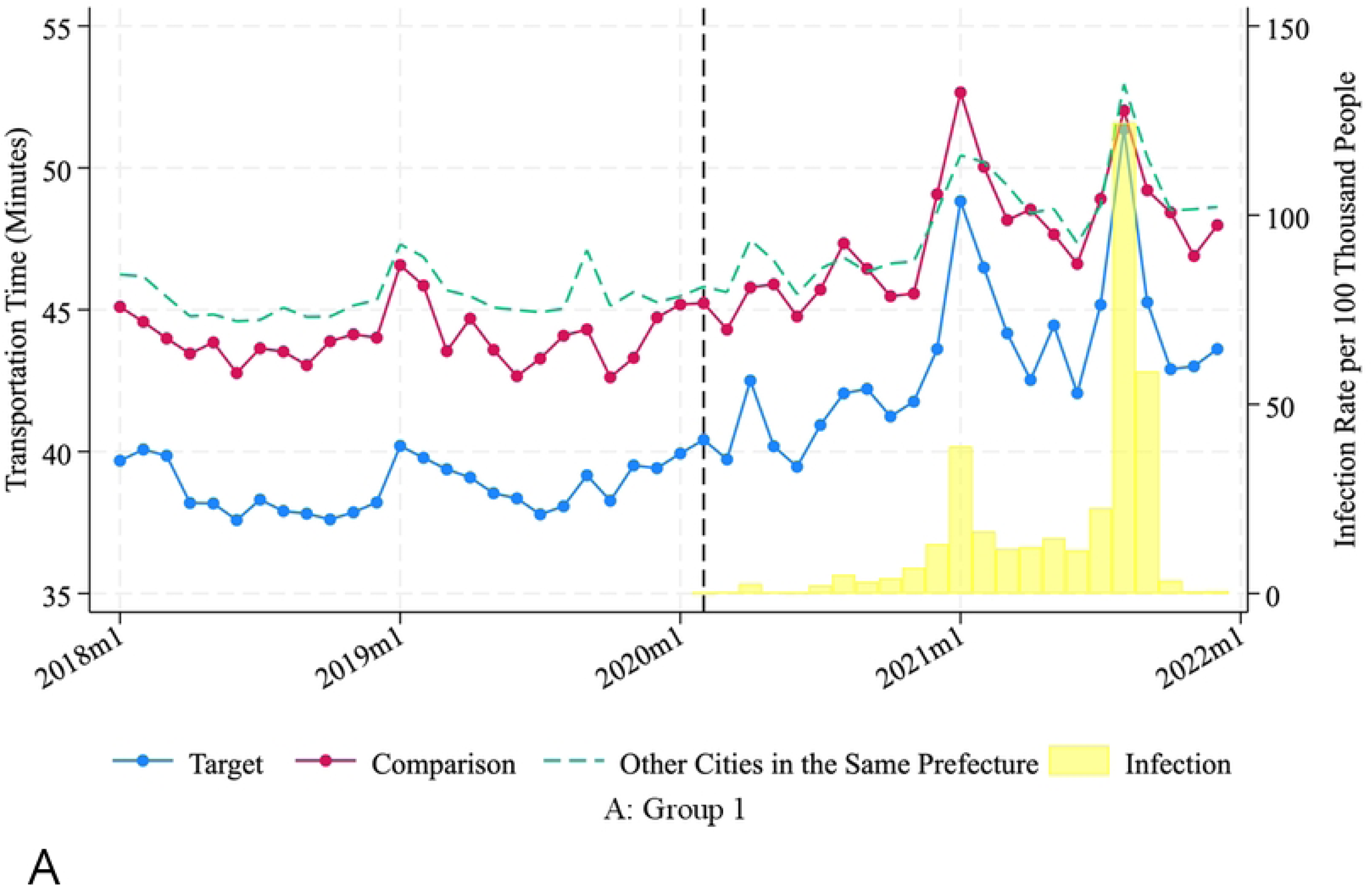
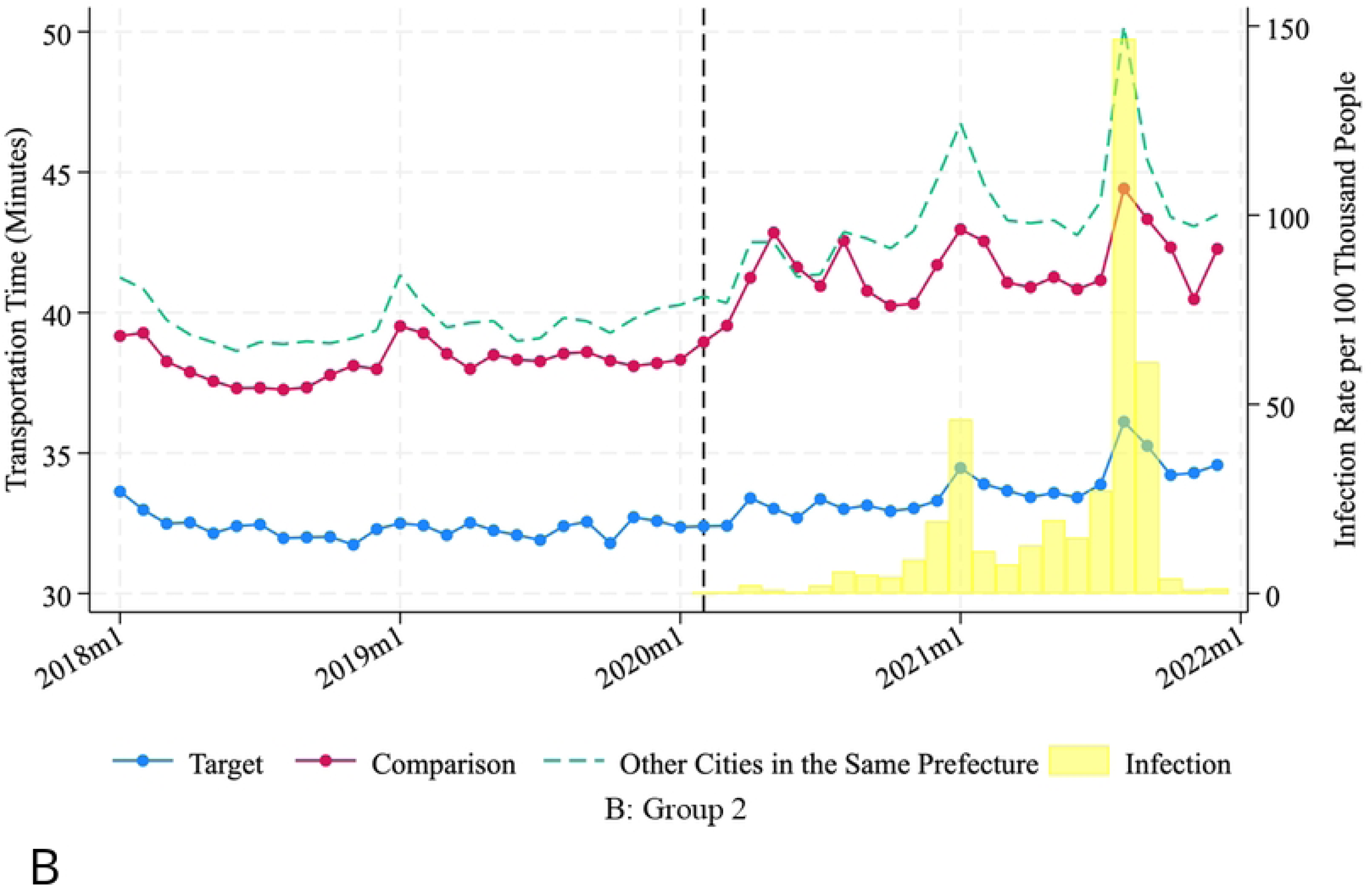
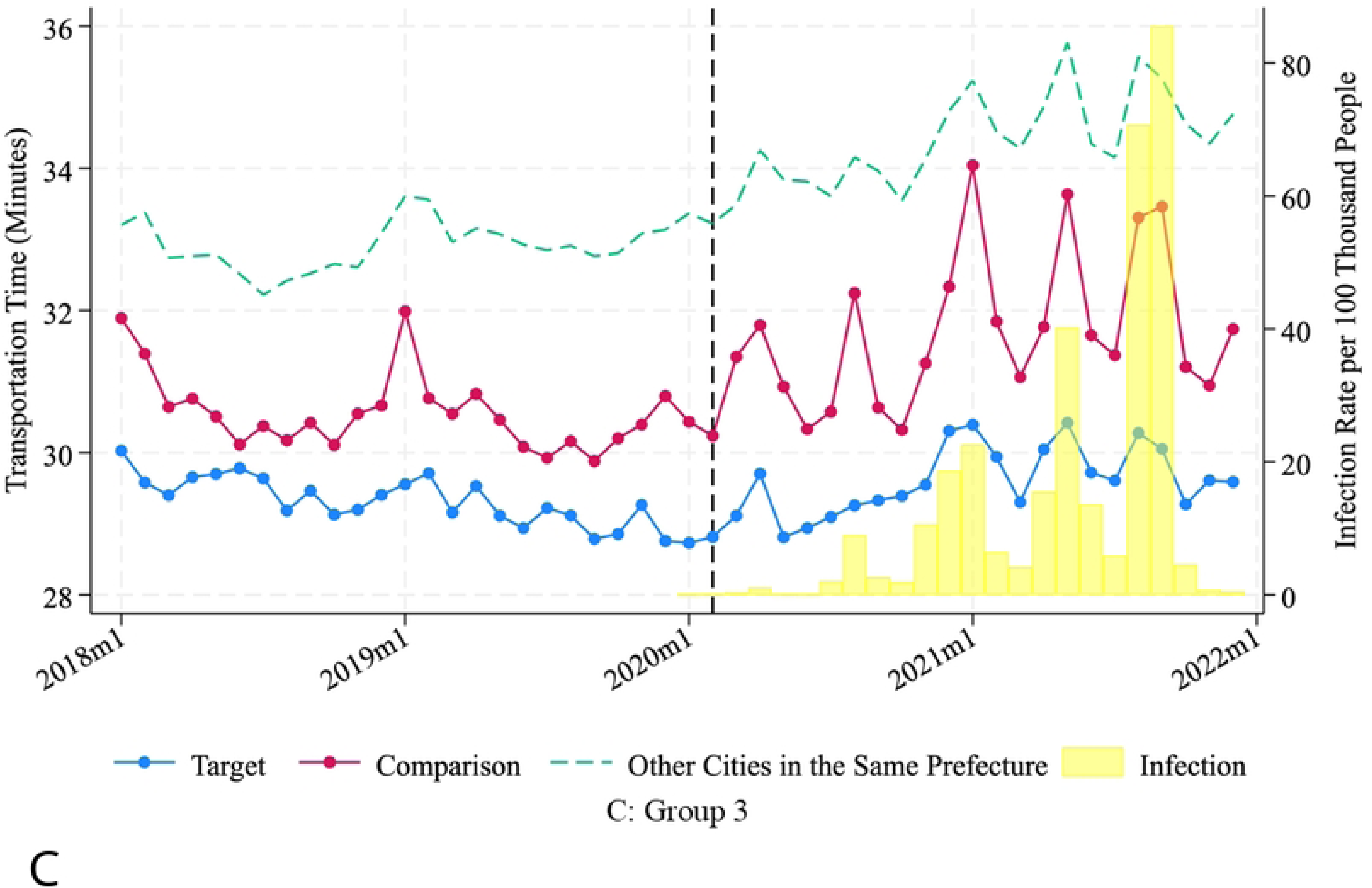
Trends of transportation time and infection rates per 100,000 populations in each area.

**Fig 3:**
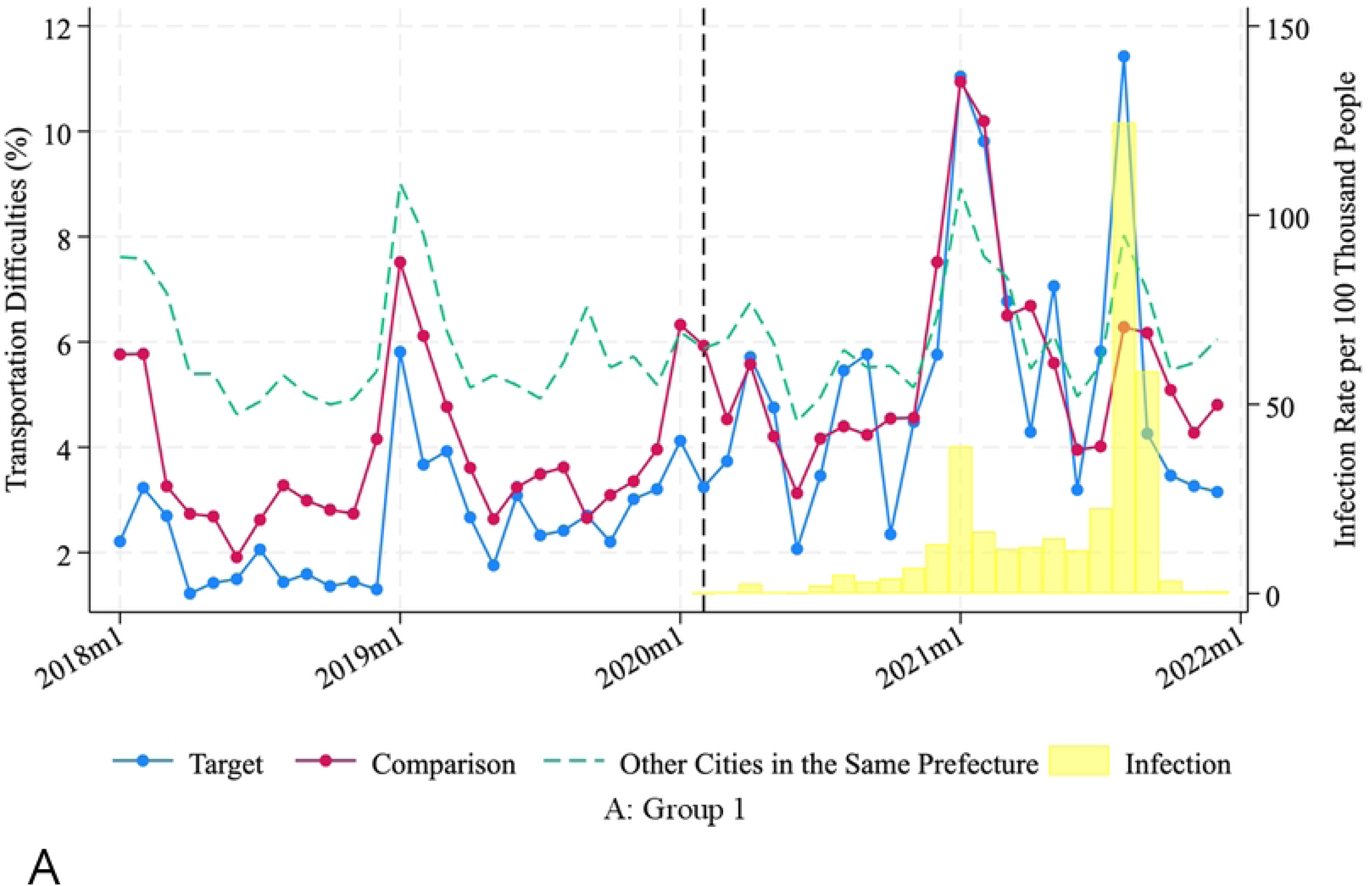
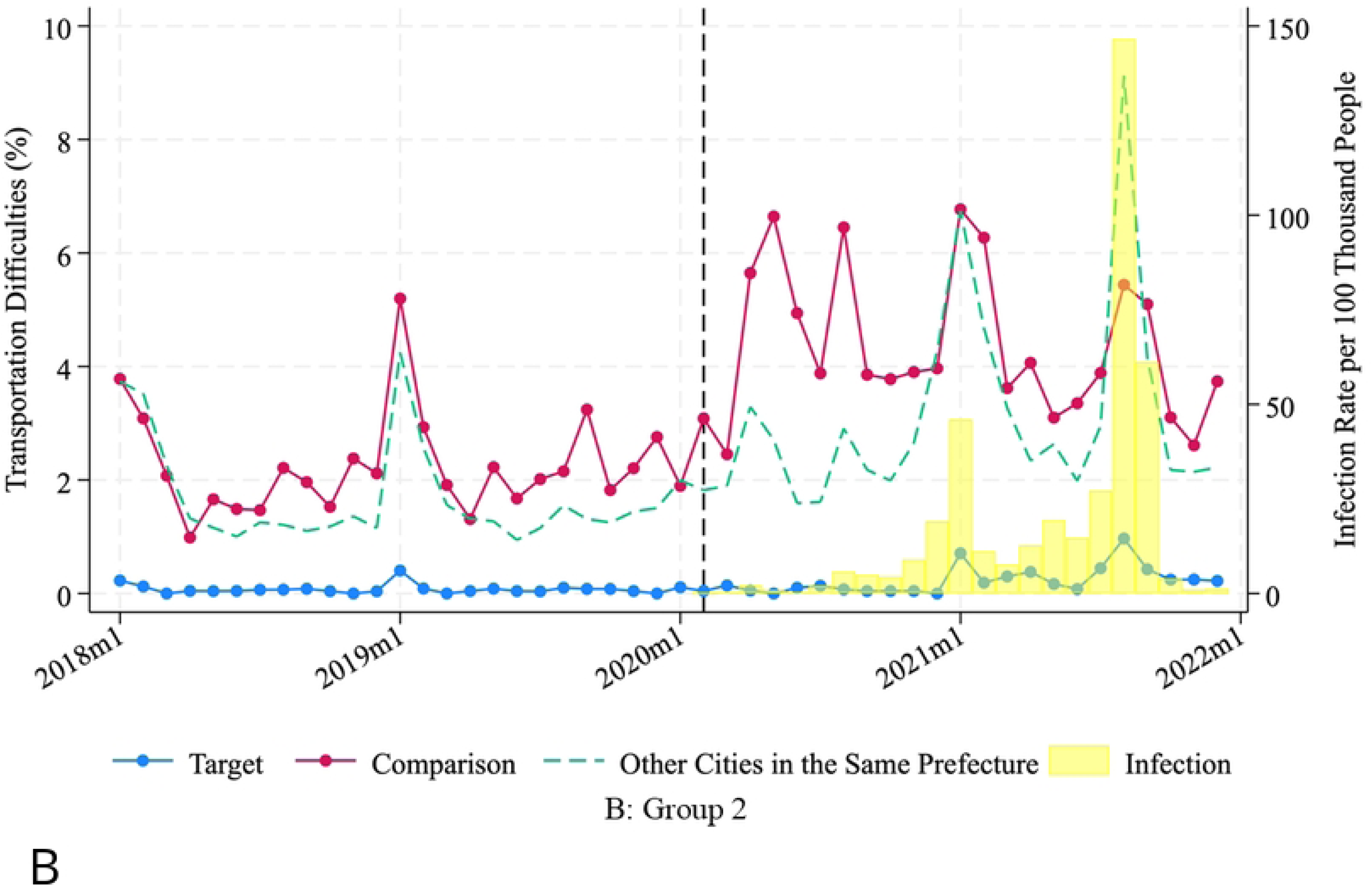
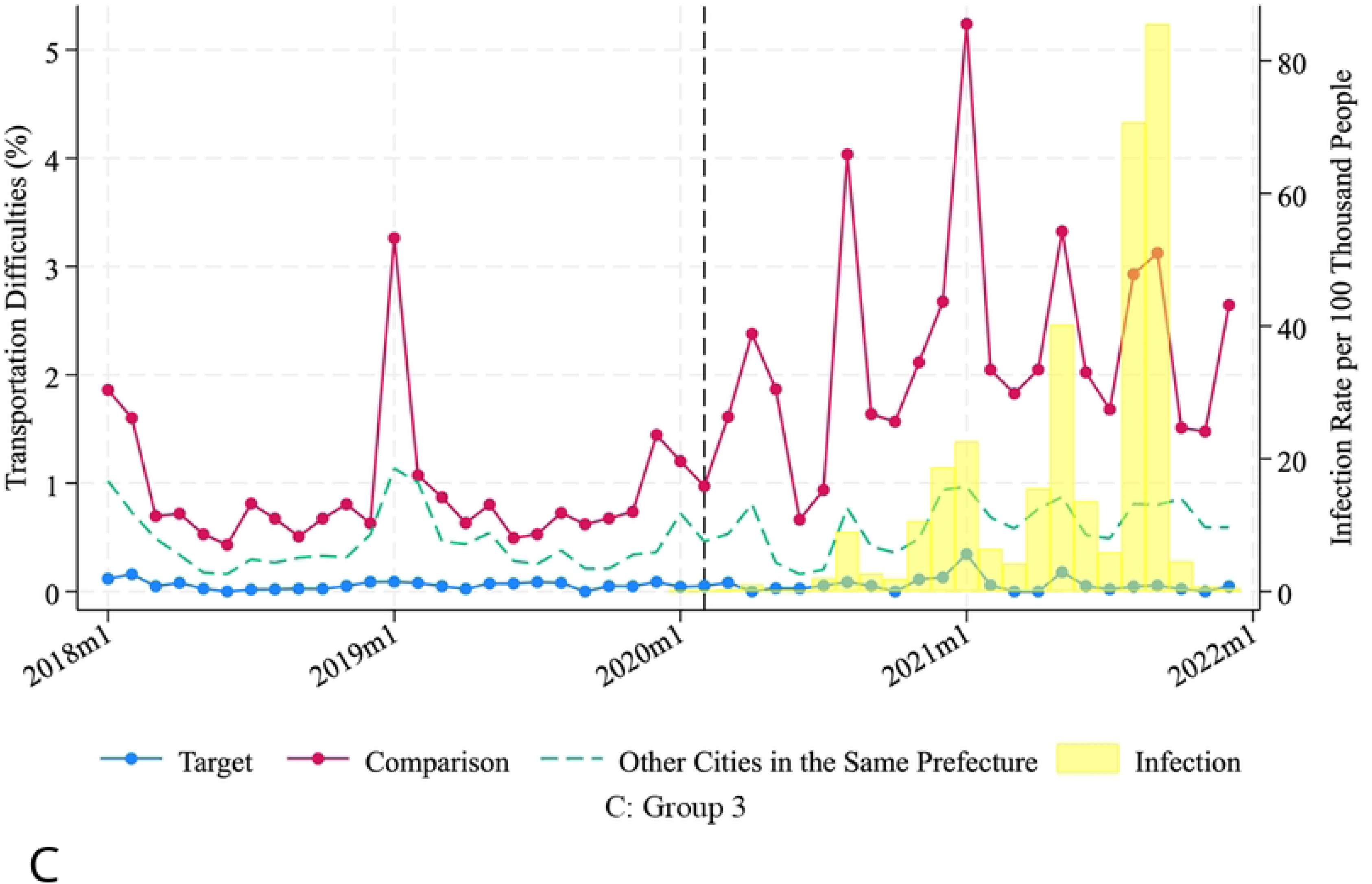
Trends of transportation difficulty cases and infection rates per 100,000 populations in each area.

**Table 3.** Multivariate regression analysis and stratified analysis of change in transportation time and number of transportation difficulty cases between before and after the first COVID-19 outbreak.

|  | Transportation time, min [Ordinary Least Squares] |  |  | Transportation difficulty, odds ratio [Logistic regression analysis] |  |  |
| --- | --- | --- | --- | --- | --- | --- |
|  | Group 1 - Chiba | Group 2 - Kanagawa | Group 3 - Aichi | Group 1 - Chiba | Group 2 - Kanagawa | Group 3 - Aichi |
| **All cases (95% CI)** | 0.214 (-1.364 to 1.792) | -2.016 (-3.631 to -0.400) | -0.606 (-1.114 to -0.098) | 1.393 (1.314 to 1.477) | 0.131 (0.110 to 0.157) | 0.086 (0.066 to 0.112) |
| Mean of Dependent Variable | 43.28 | 35.45 | 30.45 | 0.042 | 0.013 | 0.010 |
| Observations | 150856 | 186965 | 516656 | 150856 | 186965 | 516656 |
| **Respiratory diseases (95% CI)** | 3.378 (1.265 to 5.492) | -4.010 (-6.952 to -1.067) | -1.141 (-1.791 to -0.491) | 2.767 (2.349 to 3.260) | 0.105 (0.059 to 0.185) | 0.075 (0.031 to 0.182) |
| Mean of Dependent Variable | 42.27 | 34.96 | 30.36 | 0.034 | 0.013 | 0.010 |
| Observations | 13454 | 13640 | 50636 | 13454 | 12940 | 50428 |
| **Demand for oxygen (95% CI)** | 2.350 (-0.144 to 4.845) | -2.251 (-4.123 to -0.379) | -0.948 (-1.487 to -0.409) | 1.829 (1.630 to 2.052) | 0.180 (0.128 to 0.254) | 0.119 (0.073 to 0.194) |
| Mean of Dependent Variable | 44.28 | 35.6 | 30.33 | 0.065 | 0.014 | 0.009 |
| Observations | 23697 | 39040 | 108028 | 23697 | 36986 | 108028 |
| **Severe (95% CI)** | 1.573 (-0.798 to 3.945) | -1.440 (-2.959 to 0.079) | -0.419 (-0.745 to -0.092) | 1.384 (1.130 to 1.696) | 0.246 (0.132 to 0.457) | 0.314 (0.139 to 0.708) |
| Mean of Dependent Variable | 43.19 | 35.07 | 28.53 | 0.054 | 0.009 | 0.003 |
| Observations | 9270 | 14821 | 25691 | 9270 | 14499 | 25206 |
| **Moderate (95% CI)** | 0.751 (-1.325 to 2.828) | -2.450 (-4.332 to -0.569) | -0.971 (-1.497 to -0.445) | 1.385 (1.277 to 1.501) | 0.110 (0.086 to 0.141) | 0.088 (0.061 to 0.127) |
| Mean of Dependent Variable | 44.39 | 35.93 | 30.88 | 0.048 | 0.013 | 0.011 |
| Observations | 69278 | 101047 | 209290 | 69278 | 98028 | 209290 |
| **Mild (95% CI)** | -0.414 (-0.961 to 0.132) | -1.718 (-3.223 to -0.214) | -0.352 (-0.940 to 0.236) | 1.400 (1.274 to 1.539) | 0.156 (0.116 to 0.209) | 0.064 (0.040 to 0.100) |
| Mean of Dependent Variable | 42.27 | 34.96 | 30.36 | 0.034 | 0.013 | 0.010 |
| Observations | 71491 | 67494 | 276347 | 71491 | 67317 | 276347 |
| Month Fixed Effect | yes | yes | yes | yes | yes | yes |
| Fire Department Fixed Effect | yes | yes | yes | yes | yes | yes |
| Other Covariates | yes | yes | yes | yes | yes | yes |

Note: We used ordinary least square for transportation time and logistic regression analysis for
transportation difficulty.
Abbreviation: CI, confidence interval.

## Discussion

This retrospective observational study showed that the target areas of Group 2 and 3 showed significantly less worsening of tansportation performance during the COVID-19 pandemic compared with the comparison areas. Compared to the comparison areas, the increase in transportation time in the target areas of Group 2 and 3 was 2 min and 40 seconds shorter, respectively. In contrast, transportation time increased in the target area of Group 1 as well as in the comparison area. Even in a population limited to patients with respiratory diseases or those receiving oxygen, the target areas of Group 2 and 3 showed favorable results. To the best of our knowledge, this is the first study to analyse transportation time and difficulty during the COVID-19 pandemic in Japan compared with other cities using a large-scale database.

Few studies have been conducted on interventions to address transportation difficulties in Japan. Sato et al. reported that an increase in reimbursement for ambulance transportation acceptance was associated with decreased transportation difficulties^14^. Higashi et al. reported that the presence of DEP Model contributed to reduced pre-hospital transportation time^13^. However, these studies were conducted before the COVID-19 pandemic, and it is unknown whether these effects remain during COVID-19 pandemic. In Korea, early detection by expanded diagnostic tests and out-of-hospital treatment at therapeutic living centers are effective in countering the COVID-19 pandemic^18, 19^. In contrast, Taiwan successfully controlled COVID-19 by non-pharmaceutical interventions including finding, testing, tracing, isolating, and supporting (FTTIS) COVID-19 cases^20^. However, no studies have verified the effectiveness of these countermeasures statistically. In this study, we discuss the interventions that can contribute to favorable transportation times and difficulties by comparing transportation results between areas with and without a DEP Model in Japan.

We speculated several reasons why target areas of Groups 2 and 3 outperformed nearby areas, although this phenomenon was not observed in Group 1. First, Table 1 shows that large-scale hospitals are densely packed in the target areas of Groups 2 and 3 compared to Group 1. This could be attributed to the favorable results in Groups 2 and 3; namely, the small-scale ER could not function effectively during the pandemic, and the importance of large-scale ER might become apparent during the pandemic.

Shonan Kamakura General Hospital in Group 2 target area built a makeshift building with 180 beds immediately next to the hospital in May 2020 to solve the shortage of beds due to the COVID-19 pandemic. Similarly, the Ichinomiyanishi Hospital in Group 3 target area built a makeshift building with 25 beds immediately next to the hospital. These additional beds might have contributed to the favorable transportation results in Groups 2 and 3.

Despite the development of vaccines and treatments for COVID-19, epidemics of COVID-19 still occur periodically. Furthermore, it is possible that new infectious diseases with high infectivity will emerge and cause future pandemics. Then, we will face the pre-hospital transportation problems again. Based on this study, a temporary facility that accommodates patients with the pandemic disease just next to a large-scale hospital, and merging small-scale hospitals to large-scale hospitals may be beneficial in preparation for the next pandemic.

Our study had some limitations. First, although we tried to minimize the bias caused by regional variations in COVID-19 prevalence, there remain concerns that such variations may have affected the outcomes. Second, the data lacked post-transportation information. Thus, it remains unclear whether changes in transportation outcomes, such as a 40-second reduction in transportation time, have a significant impact on patient outcomes. Third, we only analysed pre-hospital data in Japan, and the results may not be generalizable to other countries where pre-hospital settings are different. Fourth, this was a retrospective observational study, and we could not determine causation.

## Conclusion

The DEP Model alone was not significantly associated with favorable transportation outcomes during the pandemic. Large-scale hospitals and makeshift buildings may be necessary to improve the transportation outcome during pandemic.

## Acknowledgements

We are thankful to all the emergency medical service personnel and healthcare workers who treated the patients and volunteered to collect the data.

## Author Contributions

**Conceptualization**: Kaiho Hirata, Takashi Shiga.

**Data curation**: Reo Takaku, Chen Meilai.

**Formal analysis**: Reo Takaku, Chen Meilai.

**Investigation**: Reo Takaku, Chen Meilai.

**Methodology**: Kaiho Hirata, Takuyo Chiba, Reo Takaku, Chen Meilai, Takashi Shiga.

**Project administration**: Kaiho Hirata.

**Supervision**: Shunya Ikeda, Takashi Shiga.

**Visualization**: Reo Takaku, Chen Meilai.

**Writing– original draft**: Kaiho Hirata.

**Writing– review & editing**: Takuyo Chiba, Reo Takaku, Takashi Shiga.

## Patient consent for publication

Not applicable.

## Ethics approval

This study was approved by the Ethics Committee of the International University of Health and Welfare, Narita Hospital (24-Im-014 June 25, 2024). Informed consent was not required to conduct this study because it was an observational study without intervention, and the data did not include identifiable personal information.

## Data availability

The database is accessible only to those who have received approval from the Japanese government. The data access is restricted based on consent agreements and Institutional Review Board approvals, permitting external researchers to use the data for research monitoring. Ownership of the data resides with the Japanese government. Researchers can contact the Ministry of Internal Affairs and Communications, Fire and Disaster Management Agency, Ambulance Service Planning Office at +81-3-5253-7529 for further information (https://www.fdma.go.jp/about/access/access002.html).

## Supporting information

S1 Figure (a) Geographical relationship between the target and comparison areas in groups 1 and 2. (b) Geographical relationship between target and comparison areas in group 3.

Note: The colors represent the percentage of cases in which a destination was determined after one call in 2021.

S2 Table. Full estimation results for main analysis

